## Supplementary Material for "Cost and cost-effectiveness of Hepatitis C virus self-testing in four settings: an economic evaluation"

### Case study assumptions and current testing guidelines in each setting

The current guidelines for testing and treatment for HCV vary in each of the four country settings.

In Georgia, an ongoing HCV elimination program that began in 2015 involves a large scale up in access to testing and treatment at very low cost to the patient[1]. A sero-survey conducted in 2021 indicates that middle aged men still hold a high burden of infections and have been accessing treatment at a lower rate than other groups (data pending publication). The standard of care is that HCV testing is widely available, including mandatory testing for all inpatients. We assume that self-testing will be implemented on a postal model, with self-tests distributed through the post following outreach to the target population of men aged 40-49. This would not be integrated within any existing HIV self-testing program.

In Kenya, we focus on PWID, where the standard of care is to have drop-in harm reduction centers which offer facility-based HCV testing[2]. National guidelines recommend regular testing of PWID and other high risk groups[3]. The self-testing approach would be peer-led testing, whereby outreach workers from the harm reduction centers go into the community with self-tests and provide a demonstration and guidance on test use. We assume this would also be integrated within existing HIV self-testing programs, as costs are based on adding HCVST to HIVST in other east African settings[4].

In Vietnam we also assume that PWID would be tested through a peer-led testing approach. Ongoing research uses respondent-driven sampling surveys to recruit PWID to testing and treatment interventions[5]. Like in Kenya, we assume that outreach workers from community-based organisations that provide harm reduction services would provide the target population with demonstration and guidance on self-test usage. However, we assume this would be separate from any HIV self-testing programs.

In China, where the target population is MSM, we assume that existing testing is available privately or through community-based organisations. The model of care for self-testing would be advertising through social media and posting tests when requested. An existing study of HIV self-testing used this model and the cost of the test was reimbursed to the patient if the test result was uploaded, we assume the HCVST program would build on this model[6].

### Supplementary Figures

#### Supplementary Figure 1

*The number of people diagnosed in each setting for each modelled scenario considered in the sensitivity analysis, compared to the counterfactual with no HCVST (in green), and the base case with HCVST (in red). Bars outlined in black indicate the scenarios with and without HCVST in which EIA is the standard of care antibody test.*

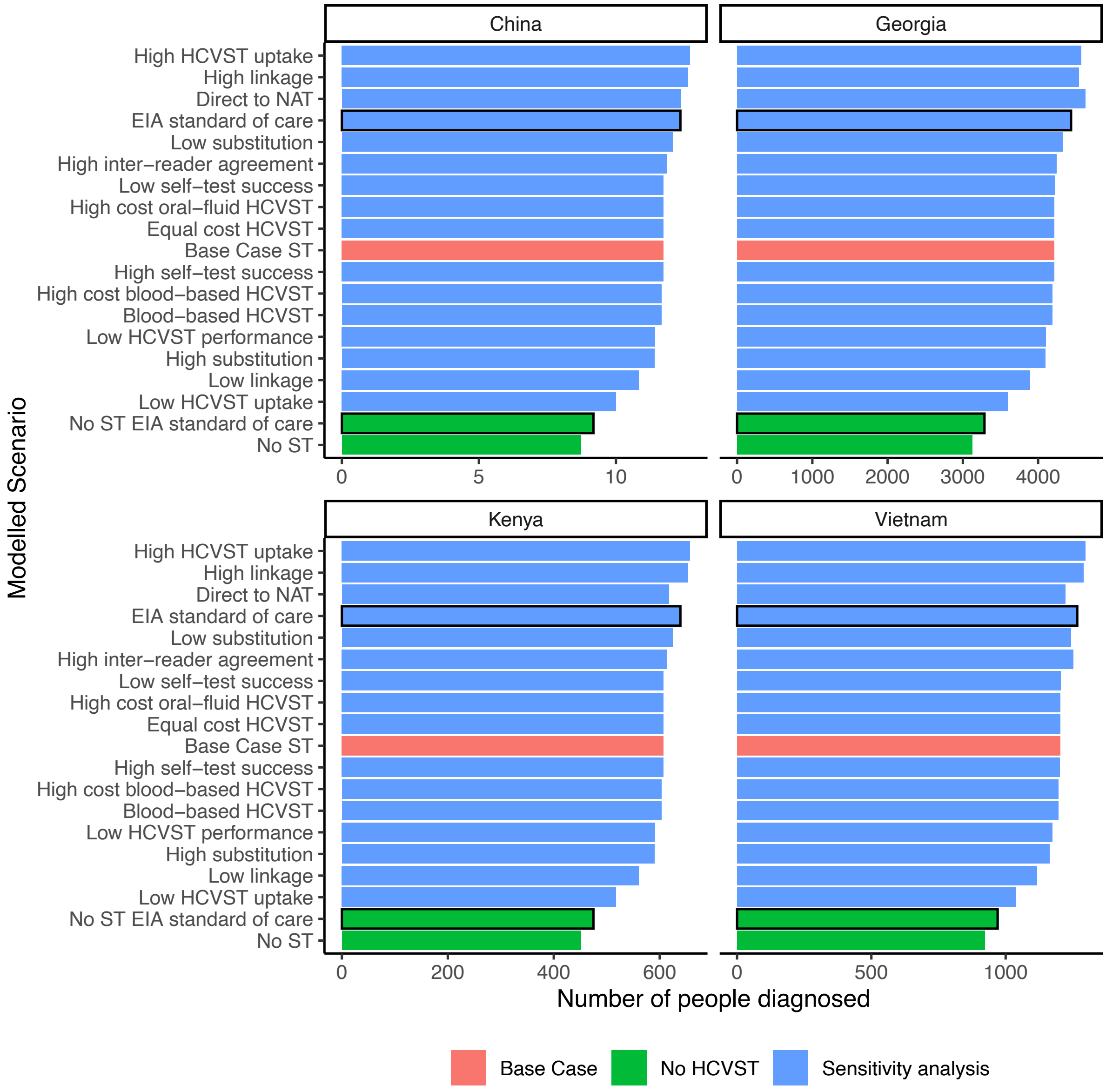

Supplementary Figure 2

*The cost of implementing HCVST per diagnosed patient (excluding the costs of treatment) in each population, for each modelled sensitivity analysis, compared to the counterfactual with no HCVST (in green), and the base case (in red). Bars outlined in black indicate the scenarios with and without HCVST in which EIA is the standard of care antibody test.*

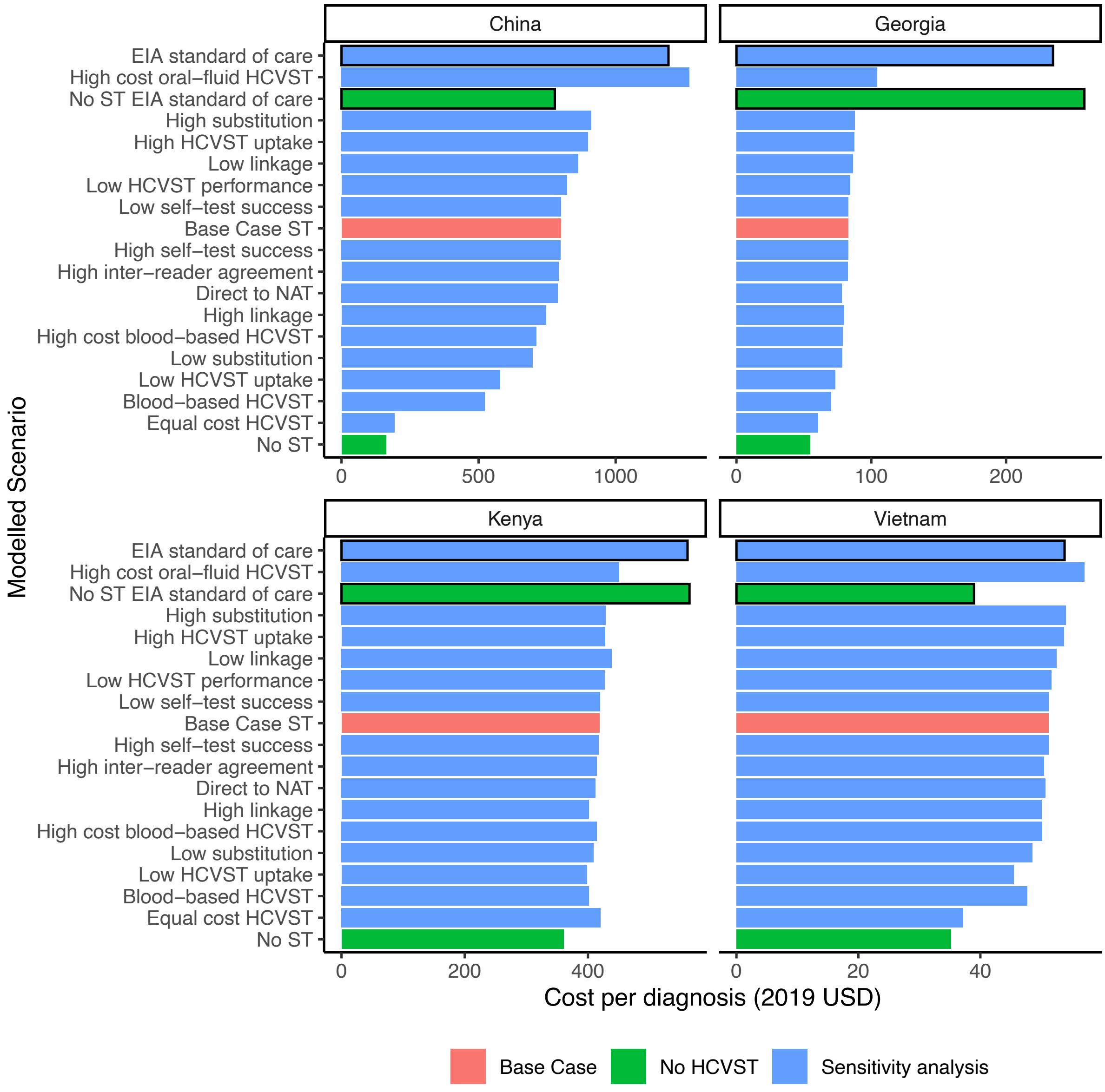

### Supplementary Figure 3

*Tornado plot showing the impact of varying parameters in sensitivity analysis on the incremental cost per cure. Note that the x-axis scale is different for each country.*

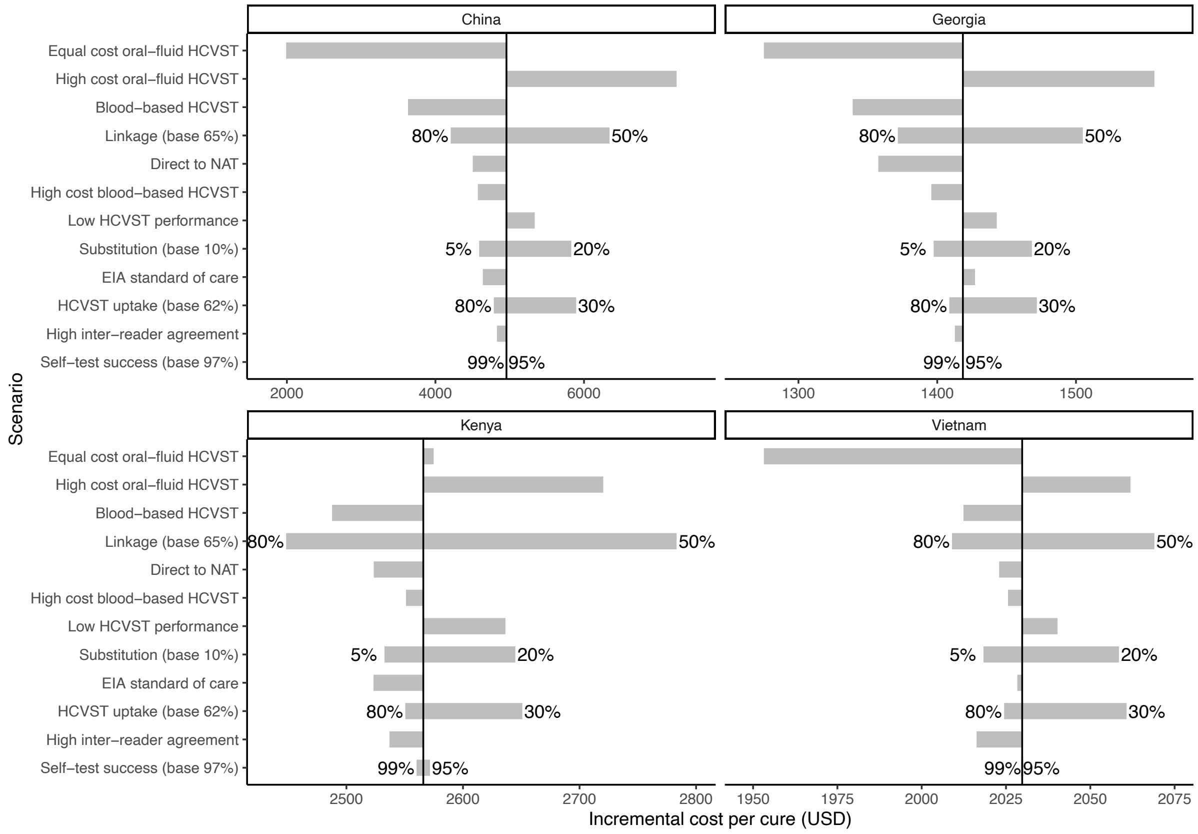

*The vertical line represents the base case incremental cost per cure as shown in Table 6, and the end of each bar represents the incremental cost per cure in each modelled scenario, with the length of the bar representing the magnitude of the difference from the base case.

### Supplementary Tables

#### Supplementary Table 1

*Transition and cost parameters used in model represented in Figure 1. At each step, the transition parameters leaving a particular cell sum to 1.*

| Step | Transition to step | Cost at step |
|---|---|---|
| Initial population | Total population of interest * proportion with unknown status | - |
| ***Of initial population:*** | | |
| Proportion receiving facility-based serologic testing | Standard testing rate minus proportion that use self-test instead | Test cost |
| Proportion receiving self-testing | New tests plus proportion that switch to using self-test instead of standard test | Self test cost + distribution |
| Proportion not tested | 1 – standard testing and self-testing | - |
| ***Of those with facility-based serologic testing:*** | | |
| Anti-HCV positive by facility-based (FB) test | Prevalence * (FB sensitivity) + (1-prevalence)*(1- FB specificity) | - |
| Anti-HCV negative by facility-based (FB) test | 1 – [Prevalence * (FB sensitivity) + (1-prevalence)*(1-FB specificity)] | - |
| ***Of those using self-testing:*** | | |
| HCVAb positive by self-test (ST) accessing facility (direct to NAT or standard care pathway) | (prevalence*ST sensitivity*inter-reader agreement + (1-prevalence)*(1-ST specificity*inter-reader agreement)) * (1 - % test failure)* % link to care if positive | - |
| HCVAb negative by self-test (ST) accessing facility | (1 – (prevalence*ST sensitivity*inter-reader agreement + (1-prevalence)*(1-ST specificity*inter-reader agreement)) * (1 - % test failure) * % link to care if negative | - |
| Unclear result or self-test failure (invalid) | % test failure * % link to care if invalid | - |
| Self-test result not reported | 1 – total above three rows | - |
| ***Of those reporting self-test results (repeat serologic testing scenario):*** | | |
| Retest antibody after invalid HCVAb self-test | % re-tested if link to care | Test cost |
| Retest antibody (repeat serologic testing scenario) after positive HCVAb self-test | % re-tested if link to care | Test cost |
| Don't retest antibody after self test | 1 – % re-tested antibody | - |
| ***Of those receiving facility-based serologic testing after positive self-test:*** | | |
| HCVAb positive by facility-based (FB) test after positive self-test (ST) | [(Prevalence * ST sensitivity*inter-reader agreement) * FB sensitivity + (1 - Prevalence)* (1-ST specificity*inter-reader agreement) * (1-FB specificity)] / [Prevalence * (ST sensitivity*inter-reader agreement) + (1-prevalence)*(1-ST specificity*inter-reader agreement)] | - |
| HCVAb negative by facility-based after positive self test | 1 – positive by facility-based test | - |
| ***Of those receiving facility-based serologic testing after invalid self-test:*** | | |
| HCVAb positive by facility-based serologic test after invalid self test | Prevalence * (FB sensitivity) + (1-prevalence)*(1- FB specificity) | - |
| HCVAb negative by facility-based serologic after invalid self test | 1 - Prevalence * (FB sensitivity) + (1-prevalence)*(1-FB specificity) | - |

Supplementary Table 2

*Transitions and costs from confirmation of viraemic infection onwards (shown in Figure 2).*

| Step | Transition to step | Cost at step |
|---|---|---|
| ***Of those anti-HCV positive eligible for NAT testing:*** | | |
| Confirm infection by NAT after facility-based test | % Receive NAT test | NAT cost |
| Confirm infection by NAT directly after self-test | % re-tested if linked to care | NAT cost |
| No confirmatory test | 1 – receive NAT | - |
| ***Of those receiving NAT testing:*** | | |
| Viraemic infection from self-test direct to NAT | Viraemic proportion of Ab positive * (Prevalence * (ST sensitivity*inter-reader agreement) / (Prevalence * (ST sensitivity* inter-reader agreement) + (1-prevalence)*(1-ST specificity* inter-reader agreement)) | - |
| Viraemic infection by NAT after facility-based test following self-test | Viraemic proportion of Ab positive * (Prevalence * ST sensitivity*inter-reader agreement) * FB sensitivity / (Prevalence * ST sensitivity*inter-reader agreement) * FB sensitivity + (1 - Prevalence)* (1-ST specificity*inter-reader agreement) * (1-FB specificity) | - |
| Viraemic infection by NAT after facility-based test only | Viraemic proportion of Ab positive * (Prevalence * (FB sensitivity) / (Prevalence * (FB sensitivity) + (1-prevalence)*(1-FB specificity)) | - |
| Not currently viraemic | 1 – positive NAT test | |
| ***Of those with chronic infection:*** | | |
| Link to care for pre-treatment assessment | Link to care (observed care cascade) | Pre-treatment costs |
| Not linked to care | 1 – link to care | - |
| ***Of those linked to care:*** | | |
| Start treatment | Treated (observed care cascade) | Treat cost |
| Not treated | 1 - treated | - |
| ***Of those treated:*** | | |
| SVR12 achieved | % Tested for SVR * % cure | NAT cost |
| SVR12 not achieved | % Tested for SVR * (1- % cure) | NAT cost |
| SVR12 not assessed | Not tested for SVR | - |

Supplementary Table 3

*Parameter values that are the same across all four settings*

| Parameter | Value | Source |
|---|---|---|
| Percent of facility-based tests that are replaced by self-tests | 10% | Assumption |
| Blood-based RDT sensitivity | 95% | [7] |
| Blood-based RDT specificity | 100% | [7] |
| Oral-fluid based RDT sensitivity | 98% | [8] |
| Oral-fluid based RDT specificity | 100% | [8] |
| Test failure rate (invalid result) | 3% | Assumption |
| Link to facility with positive self-test | 65% | [9] |
| Link to facility with negative self-test | 5% | Assumption |
| Link to facility with invalid self-test | 65% | [9] |
| Receive facility-based test if link to facility with positive or invalid self-test | 100% | Assumption |
| Receive follow up test in clinic if report negative self-test result | 0% | Assumption |
| Self-test unit cost (oral-fluid based) | 5.63 USD | Assumption |
| Self-test unit cost (blood-based) | 2.25 USD | Assumption |
