## Supplementary material for "Cost and cost-effectiveness of Hepatitis C virus self-testing in four settings: an economic evaluation": CHEERS checklist

### CHEERS 2022 Checklist

| Topic | No. | Item | Location where item is reported |
|---|---|---|---|
| **Title** | | | |
| | 1 | Identify the study as an economic evaluation and specify the interventions being compared. | Title: Page 1 |
| **Abstract** | | | |
| | 2 | Provide a structured summary that highlights context, key methods, results, and alternative analyses. | Abstract Page 2 |
| **Introduction** | | | |
| **Background and objectives** | 3 | Give the context for the study, the study question, and its practical relevance for decision making in policy or practice. | Introduction Page 4 |
| **Methods** | | | |
| **Health economic analysis plan** | 4 | Indicate whether a health economic analysis plan was developed and where available. | No analysis plan published |
| **Study population** | 5 | Describe characteristics of the study population (such as age range, demographics, socioeconomic, or clinical characteristics). | Not applicable |
| **Setting and location** | 6 | Provide relevant contextual information that may influence findings. | Methods page 4 and supplement page 2 |
| **Comparators** | 7 | Describe the interventions or strategies being compared and why chosen. | Methods pages 5-6 |
| **Perspective** | 8 | State the perspective(s) adopted by the study and why chosen. | Methods page 7 |
| **Time horizon** | 9 | State the time horizon for the study and why appropriate. | Methods page 5 |
| **Discount rate** | 10 | Report the discount rate(s) and reason chosen. | Not applicable |
| **Selection of outcomes** | 11 | Describe what outcomes were used as the measure(s) of benefit(s) and harm(s). | Methods page 5 |
| **Measurement of outcomes** | 12 | Describe how outcomes used to capture benefit(s) and harm(s) were measured. | Modelled |

| Topic | No. | Item | Location where item is reported |
| --- | --- | --- | --- |
| **Valuation of outcomes** | 13 | Describe the population and methods used to measure and value outcomes. | Modelled |
| **Measurement and valuation of resources and costs** | 14 | Describe how costs were valued. | Methods page 6-7 |
| **Currency, price date, and conversion** | 15 | Report the dates of the estimated resource quantities and unit costs, plus the currency and year of conversion. | Methods page 7 |
| **Rationale and description of model** | 16 | If modelling is used, describe in detail and why used. Report if the model is publicly available and where it can be accessed. | Methods page 5 |
| **Analytics and assumptions** | 17 | Describe any methods for analysing or statistically transforming data, any extrapolation methods, and approaches for validating any model used. | Not applicable |
| **Characterising heterogeneity** | 18 | Describe any methods used for estimating how the results of the study vary for subgroups. | Not applicable |
| **Characterising distributional effects** | 19 | Describe how impacts are distributed across different individuals or adjustments made to reflect priority populations. | Not applicable |
| **Characterising uncertainty** | 20 | Describe methods to characterise any sources of uncertainty in the analysis. | Sensitivity analysis Table 2 |
| **Approach to engagement with patients and others affected by the study** | 21 | Describe any approaches to engage patients or service recipients, the general public, communities, or stakeholders (such as clinicians or payers) in the design of the study. | Not applicable |
| **Results** | | | |
| **Study parameters** | 22 | Report all analytic inputs (such as values, ranges, references) including uncertainty or distributional assumptions. | Table 1 |
| **Summary of main results** | 23 | Report the mean values for the main categories of costs and outcomes of interest and summarise them in the most appropriate overall measure. | Results page 7 |
| **Effect of uncertainty** | 24 | Describe how uncertainty about analytic judgments, inputs, or projections affect findings. Report the effect of choice of discount rate and time horizon, if applicable. | Results page 8 |

| Topic | No. | Item | Location where item is reported |
|---|---|---|---|
| **Effect of engagement with patients and others affected by the study** | 25 | Report on any difference patient/service recipient, general public, community, or stakeholder involvement made to the approach or findings of the study | Not applicable |
| **Discussion** | | | |
| **Study findings, limitations, generalisability, and current knowledge** | 26 | Report key findings, limitations, ethical or equity considerations not captured, and how these could affect patients, policy, or practice. | Discussion page 8-9 |
| **Other relevant information** | | | |
| **Source of funding** | 27 | Describe how the study was funded and any role of the funder in the identification, design, conduct, and reporting of the analysis | Title page (1) |
| **Conflicts of interest** | 28 | Report authors conflicts of interest according to journal or International Committee of Medical Journal Editors requirements. | Title page (1) |

*From:* Husereau D, Drummond M, Augustovski F, et al. Consolidated Health Economic Evaluation Reporting Standards 2022 (CHEERS 2022) Explanation and Elaboration: A Report of the ISPOR CHEERS II Good Practices Task Force. Value Health 2022;25. doi:10.1016/j.jval.2021.10.008
